# The prevalence and factors associated with premature birth among post-delivery women in the Mbeya region: An analytical cross-sectional study

**DOI:** 10.1101/2022.11.14.22282321

**Authors:** Glory Godfrey Mawolle, Fabiola Vincent Moshi

## Abstract

In Tanzania, there was an increase of prematurity rate from 11.4% in 2014 up to 16.60% in 2016 (1). This is a hospital based analytical cross-sectional study which involved biopsychosocial model, which focused on identifying prevalence and associated factors for preterm births among post-delivery women in Mbeya region, one of Tanzania regions. This study involved hospitals in Mbeya urban, Mbeya rural, Chunya, Kyela, Mbarali, Rungwe, Busokelo and Tukuyu districts, where the prevalence of preterm births in Mbeya found to be 39.1%. The study pointed out that factors associated with preterm births were child spacing of <24months (AOR=3.058; 95% CI = 1.026-9.116: p-value 0.045), non-effective use of malaria prophylaxis during pregnancy (AOR=5.418; 95% CI =1. p-value 0.008), twin pregnancy (AOR=4.657; 95% CI =2.112-10.223, p-value < 0.001), violence during pregnancy (AOR=2.059; p-value 0.048), lack of social support (AOR=1.993; p-value 0.022) and use of pica during pregnancy (AOR=1.880; p-value 0.029).

The study outcome revealed that the prevalence of preterm births in Mbeya Region is even higher. Therefore, to minimize or eliminate the problem a deliberate effort to come up with strategies to improve family planning, applications of antimalaria prophylaxis, stop the use of pica and violence during pregnancy was highly recommended.

## Introduction

Premature birth has recently gained global attention due to the adverse impact if not well addressed. Of 15million premature births in a year globally, over 84% occur at 32-36 weeks of gestation and categorized as late preterm. About 5% only fall into the extremely preterm birth of less than 28 weeks category and the other 10% births occur at 28-32 weeks of gestation and categorized as very preterm (1).

Preterm birth rates are estimated to be 11.1% worldwide of which South Asia and Sub-Saharan Africa accounts for 60% of all premature births (2). Low-income countries have an average of 12.2% of preterm birth whereas 9.4% and 9.3% correspondingly for middle and higher-income countries (3). Tanzania a sub-Saharan country has been reported to be among the top 10 countries with the highest number of preterm babies in which 336,000 babies are born too early each year (2). In Tanzania, preterm is regarded to be the primary factor in one out of every four newborn deaths, while prematurity-related conditions are thought to account for 23% of all newborn deaths (8).

The Mbeya Region in Tanzania where this study focused in determining the true prevalence and factors of preterm delivery had 31% of neonatal deaths caused by prematurity (16). Little was known about the prevalence and factors associated with premature birth in this region. Therefore, this study aimed to identify prevalence and factors associated with preterm birth in Mbeya Region, Tanzania.

The literature indicates that globally, an estimated 15 million babies are born too early every year which is more than 1 in 10 babies (3). Premature births account for 11.1% of the world’s live births, ranging from 5% in European countries and 18% in some of African countries, making majority of about 60% of preterm birth in South Asia and sub-Saharan Africa (2). In the poorest countries, on average, 12% of babies are born premature, compared to 9% in higher-income countries (12). Sub-Saharan Africa is the most affected in southern region with prematurity rate of 11.19% followed by eastern region with prematurity rate of 7.34% then western region with prematurity rate of 3.24% and central region with prematurity rate of 2.06% (13). The major possible causes of the problem have been stated to be infections, chronic maternal illness such as hypertension and diabetic mellitus and genetic inspirational, a prior history of preterm birth, underweight, obesity, diabetes, hypertension, smoking, infection, maternal age of either under 17 years or over 40 years, genetics, multi-fetal pregnancy either twins, triplets, or higher and pregnancies spaced too closely together (1).

Through this study, the objective was achieved indicating the preterm births in Mbeya region to be 39.1% and associated factors being child spacing, poor application of malaria prophylaxis, twin pregnancies, violence, lack of social support and use of pica during pregnancy.

## Material and Methods

A census method was used to select all district and referral hospitals and a simple random sampling with lottery replacement technique was used to select study participants. A structured questionnaire was used for data collection. Frequencies, percentages, chi-square analysis, and binary logistic regression methods were used for data analysis using SPSS version 25 to compare and explain the prevalence and preterm delivery contributing variables. A variable in the final model was judged statistically significant when it had a 95% Confidence Interval (CI) and a p-value of 0.05.

### Study design and setting

This study was an analytical cross-sectional conducted from April to May 2022.The study was conducted in Mbeya region of Tanzania. The region have 5 districts with 6 local Government Authorities (17). The Region has a population of 2,707,410 (17). The region’s 2.7 percent average annual population growth rate was tied for the tenth highest in the country. The Region also had 557,574 number of women of reproductive age in 2021 with fertility rate of 130 births/1000 women (18). More over Mbeya Region contain 1 Zonal referral hospital, 1 Regional Referral Hospital, 6 Districts hospitals, 25 health centers and 270 dispensaries (17). All district hospitals and referral hospitals have neonatal units and maternity wards which provide care to both mothers and their newborns including those who are premature.

### Study Population

This study involved all postnatal mothers in selected public health facilities in Mbeya Region, with live births and their neonates within 24 hours of giving birth, and those who had consent to participate. This study did not involve postnatal mothers who had stillbirths and those who did not give consent to participate. Also, postnatal mothers who were extremely ill or whose infants were seriously ill and unsuitable for interviews were not included.

### Sample size and sampling procedure

Estimated minimum sample size (N) for this study was calculated by using the formula;

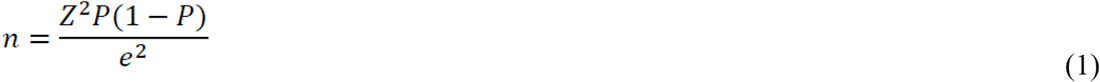

Whereby, n = Minimum sample size, Z = Critical value (1.96) at 95% confidence level, e = Margin of error 5% (type 1 error) for prevalence between 10% and 90% P = Sample proportion of premature rate in Tanzania 16.6% (1).

### Selection of participants

All post-delivery who gave birth within 24 hours were included in the study. A proportionate sample was computed to get sample size in each hospital. Finally, a simple random sampling with lottery replacement method was applied to obtain participants of the study.

Proportionate from each group = (Sample size x population in each hospital) / Total deliveries

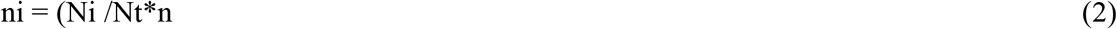

Whereby.

ni = Required number of delivered from hospitals

Ni = Total number of deliveries in hospital in a month

Nt = Total number of deliveries in all hospitals

n = estimated sample size

## Results

The prevalence of preterm births in Mbeya Region was found to be 39.1%. Maternal factors associated with preterm birth were child spacing <24months (AOR=3.058; 95% CI = 1.026-9.116: p-value 0.045) and poor applications of malaria prophylaxis (AOR=5.418; 95% CI =1. p-value 0.008) (table 1). Neonatal factor associated with prematurity was twin pregnancy (AOR=4.657; 95% CI =2.112-10.223, p-value < 0.001) (table 2). Violence during pregnancy (AOR=2.059; p-value 0.048), lack of social support (AOR=1.993; p-value 0.022) and use of pica during pregnancy (AOR=1.880; p-value 0.029) were Social behavioral factors associated with prevalence of prematurity (table 3)

**Table 1.**
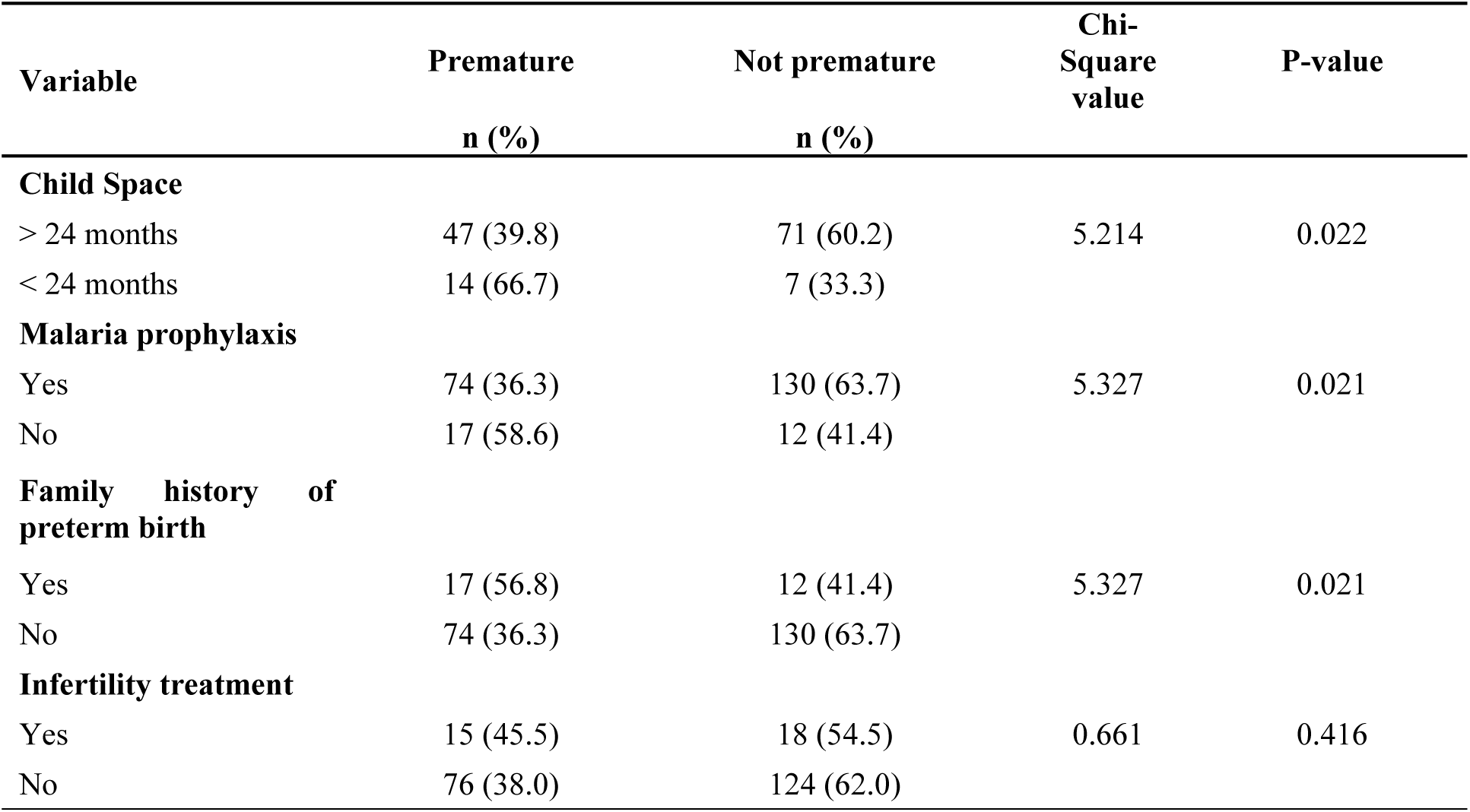
Obstetric factors associated with preterm births. Child spacing <24months (AOR=3.058; 95% CI = 1.026-9.116: p-value 0.045) and poor applications of malaria prophylaxis (AOR=5.418; 95% CI =1. p-value 0.008).

| Variable | Premature<br>n (%) | Not premature<br>n (%) | Chi-Square<br>value | P-value |
| --- | --- | --- | --- | --- |
| **Child Space** |  |  |  |  |
| > 24 months | 47 (39.8) | 71 (60.2) | 5.214 | 0.022 |
| < 24 months | 14 (66.7) | 7 (33.3) |  |  |
| **Malaria prophylaxis** |  |  |  |  |
| Yes | 74 (36.3) | 130 (63.7) | 5.327 | 0.021 |
| No | 17 (58.6) | 12 (41.4) |  |  |
| **Family history of preterm birth** |  |  |  |  |
| Yes | 17 (56.8) | 12 (41.4) | 5.327 | 0.021 |
| No | 74 (36.3) | 130 (63.7) |  |  |
| **Infertility treatment** |  |  |  |  |
| Yes | 15 (45.5) | 18 (54.5) | 0.661 | 0.416 |
| No | 76 (38.0) | 124 (62.0) |  |  |

**Table 2.** Neonatal factor associated with preterm births. Neonatal factor associated with prematurity was twin pregnancy (AOR=4.657; 95% CI =2.112-10.223, p-value < 0.001).

| Variable | Premature<br>n (%) | Not premature<br>n (%) | Chi-Square value | P-value |
| --- | --- | --- | --- | --- |
| **Pregnancy outcome** |  |  |  |  |
| Single fetus | 63 (33.5) | 125 (66.5) | 12.575 | <0.001 |
| Twins | 28 (62.2) | 17 (37.8) |  |  |
| **Fetal distress** |  |  |  |  |
| Yes | 12 (48.0) | 13 (52.0) | 0.941 | 0.332 |
| No | 79 (38.0) | 129 (62.0) |  |  |
| **Congenital fetal Anomalies** |  |  |  |  |
| Yes | 1(50) | 1(50) |  | 1.000 |
| No | 141(61.0) | 90(39.0) |  |  |
| **APGARSCORE (1 -5 minutes)** |  |  |  |  |
| 4-6 | 17 (56.7) | 13(43.3) | 4.487 | **0.034** |
| 7-10 | 74 (36.5) | 129(63.5) |  |  |
| **Birth weight (g)** |  |  |  |  |
| < 2500 | 81 (94.2) | 5 (5.8) | 174.060 | <0.001 |
| > 2500 | 10 (6.8) | 137 (93.2) |  |  |

**Table 3.** Social behavioral factors/characteristics. Violence during pregnancy (AOR=2.059; p-value 0.048), lack of social support (AOR=1.993; p-value 0.022) and use of pica during pregnancy (AOR=1.880; p-value 0.029)

| Variable | Frequency (n) | Percentage (%) |
| --- | --- | --- |
| **Use of alcohol** |  |  |
| Yes | 21 | 9 |
| No | 212 | 91 |
| **Use of Pica** |  |  |
| Yes | 121 | 51.9 |
| No | 112 | 48.1 |
| **Supportive care** |  |  |
| Yes | 141 | 60.5 |
| No | 92 | 39.5 |
| **Planned pregnancy** |  |  |
| Yes | 147 | 63.1 |
| No | 86 | 36.9 |
| **Daily activities** |  |  |
| Full time | 50 | 21.5 |
| Part time | 183 | 78.5 |
| **Beating by spouse** |  |  |
| Yes | 22 | 9.4 |
| No | 211 | 90.6 |
| **Farming activities** |  |  |
| Yes | 77 | 33 |
| No | 156 | 67 |
| **Type of violence experienced** |  |  |
| Sexual Violence | 2 | 0.9 |
| Emotional Violence | 28 | 12 |
| Economic Violence | 5 | 2.1 |
| No violence experienced | 188 | 80.7 |

Mbeya regional hospital was leading with preterm prevalence of 52%, Mbeya zonal hospital was at second place with preterm prevalence of 50% followed by Ifisi hospital 47%, Kyera hospital 39%, Mbarali hospital 38%, Tukuyu hospital 31%, Chunya hospital 29% and Itete hospital being with lowest prevalence record of 13% (Fig.1).

**Fig 1.**
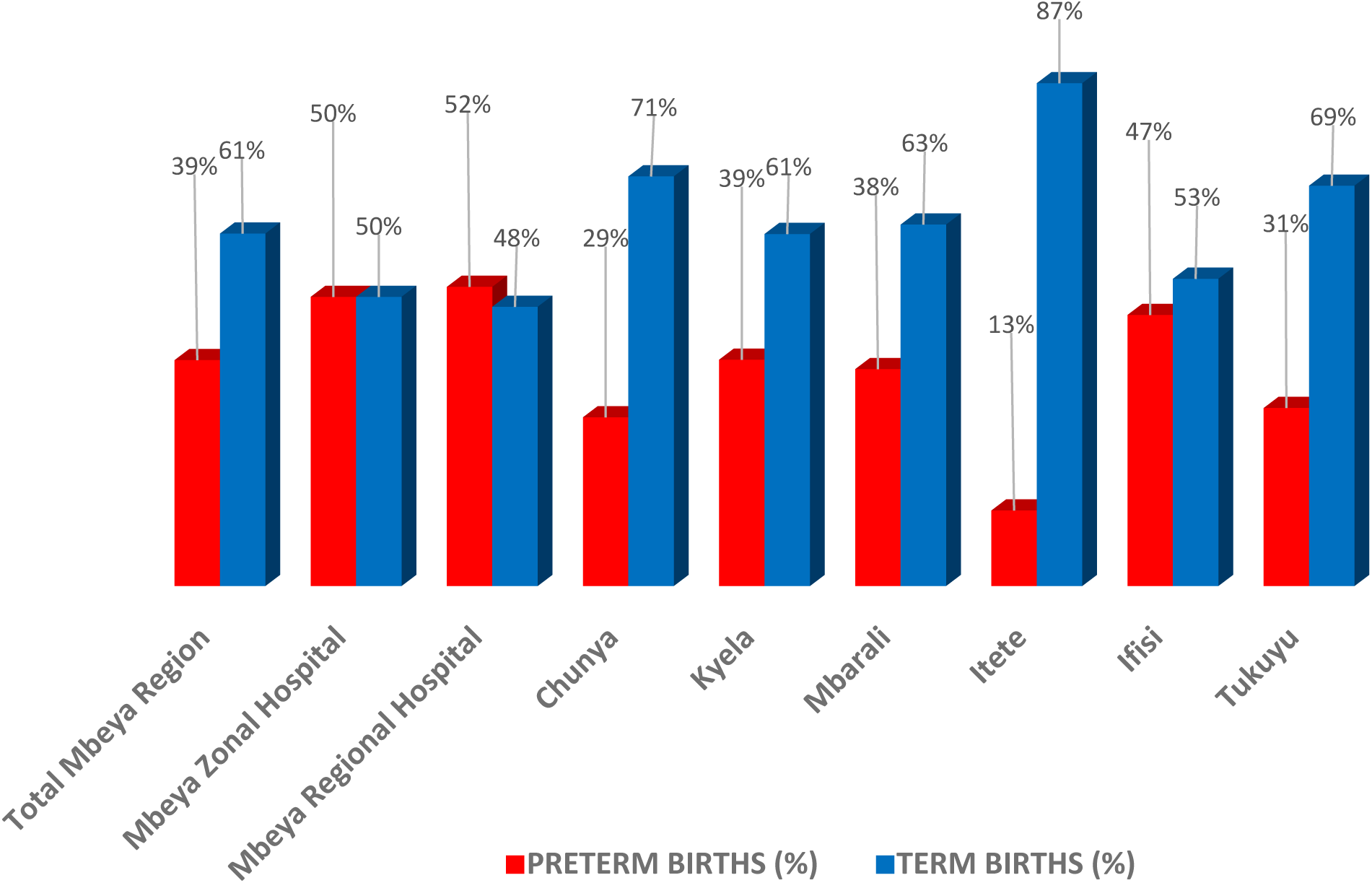
Prevalence of Preterm births in Mbeya region. Mbeya regional hospital was leading with prevalence of 52%, Mbeya zonal hospital was at second place with prevalence of 50% followed by Ifisi hospital 47%, Kyera hospital 39%, Mbarali hospital 38%, Tukuyu hospital 31%, Chunya hospital 29% and Itete hospital being with lowest prevalence record of 13%.

## Discussion

The overall prevalence of preterm birth in Mbeya region was found to be 39.1%. This is higher compared to the global rate which is 11.1% and the prevalence of preterm birth in Tanzania which is 16.6%. The elevated prevalence in this study may be due to high proportion of preterm birth which was found among participants who were not properly using malarial prophylaxis (58.6%), child space of less than 24 months (66.7%) and twin pregnancy (62.2%) of which were strongly statistically significant associated with preterm birth.

## Conclusion

The study revealed that the prevalence of preterm births in Mbeya Region is even higher. It was noted that women who are more likely to deliver preterm babies were those with less than 2years child spacing, poor adherence to anti-malaria prophylaxis, those with multiple pregnancies, those who are lacking enough social support, experiencing violence and those who are using pica during pregnancy. To ensure full term delivery, a deliberate effort to come up with strategies to improve family planning use and complete antimalarial prophylaxis is highly recommended. Public health awareness campaigns, family planning to promote ideal pregnancy spacing, and health education to the society will help to minimize/eliminate prematurity and its implications.

However, this study methodology was limited because it could not demonstrate a causal link between causes and effects, therefore it was suggested that a cohort study which is the best effective method for establishing the occurrence and natural history of illnesses to be conducted in Mbeya region to help examining numerous outcomes of a single exposure.

## Data Availability

All relevant data are within the manuscript and its Supporting Information files.

## Acknowledgements

Special acknowledgements to my supervisor, Dr. Fabiola Moshi for the advice, directions, help and support during the period of writing my dissertation, School of Nursing and Public Health at the University of Dodoma, Dr. Stephen Kibusi the Dean of the School of Nursing and Public Health and Dr. Angela Joho the Head of the Department of Clinical Nursing.

Special thanks to the Institution Review Board (IRB) of the University of Dodoma, specifically the College of Health Science, for giving their approval and ethical clearance, respectively. My sincere gratitude to the administrative authority of the Mbeya Region, Regional Administrative Secretary (RAS), and the Mbeya, Rungwe, Kyela, Mbarali, Chunya, and Busokelo District Councils for approving my requests of data collection for this study.

Finally, I want to express my gratitude to my dear husband, my family, my classmates, and my co-workers for their support during the period of my academic career.

